# Factors affecting trust in clinical trials conduct: Views of stakeholders from a qualitative study in Ghana

**DOI:** 10.1101/2022.10.05.22280221

**Authors:** Samuel Tamti Chatio, John Kuumuori Ganle, Philip Baba Adongo, Ulrike Beisel

## Abstract

**Introduction:** Globally, there are signs of declining public trust in science, especially in biomedical research. In Ghana, there are equally signs of public distrust in the conduct of biomedical research in Ghana. Typical examples are the unsuccessful conduct of the Ebola vaccine trial and the initial refusal of parents to allow their children to receive the recently piloted malaria vaccine in Ghana. Therefore, this study explored stakeholders’ views on factors affecting trust in clinical trials conduct in Ghana.

**Methods:** This was a cross-sectional exploratory study using qualitative research approach. Forty-eight in-depth interviews and Key informant interviews were conducted with stakeholders. Purposive sampling technique was used to select participants. All the interviews were recorded, transcribed and coded into main and sub-themes using QSR Nvivo 12 software to aid thematic analysis.

**Results:** Overall, participants saw the need for the conduct of clinical trials in Ghana because clinical trial studies enable scientists to come out with effective medicines for the management of diseases. Pre-implementation factors such as inadequate stakeholder engagement, rumours and negative influence affected trust. Implementation factors such as perceived risks about clinical trials medicines, apprehensions on drawing and use of blood samples, poor informed consent administration and perceived no illness all negatively affected trust in clinical trials conduct.

**Conclusion:** Trust is a fundamental factor affecting a successful conduct of clinical trials. Thus, there is need for collective efforts by all stakeholders including research institutions and clinical trial regulatory bodies to take the issue of trust in clinical trials conduct seriously.

## Introduction

Globally, there are signs of declining public trust in science, especially in biomedical research [1]. Evidence exists that scientists’ dehumanization and exploitation of people in the name of science led to suspicion and mistrust in the conduct of clinical trials [2]. The long list of unethical conduct in biomedical research including the Tuskegee syphilis study of 1932 in the United States of America; the Guatemala experiments where about 1,308 innocent people were intentionally infected with organisms of syphilis and gonorrhoea in 1946 and the Nazi doctors’ experiments subjecting prisoners to gruesome medical experiments where surgeries were done without anesthesia have all contributed to undermine public trust in biomedical research [3,2]. In Africa, there has been reported cases of public distrust in biomedical research such as the boycott of the 2003 polio vaccination campaigns in Northern Nigeria because of a number of factors, including the memory of a controversial trial of the antibiotic Trovan in 1996 and the delayed polio eradication efforts in India and Democratic Republic of Congo [4,1].

In Ghana, there are equally signs of public distrust in the conduct of biomedical research. A typical example is the Ebola Virus Disease (EVD) outbreak in West Africa, where Ghana was selected to conduct EVD vaccine trial (Adenovirus 26 vectored glycoprotein/MVA-BN (Ad26.ZEBOV/ MVA-BN) in the Volta Region was not successful because of public distrust [5,6]. Closely related to this is the initial refusal by parents to allow their children to receive the recently piloted malaria vaccine in Ghana due to lack of trust [7]. The piloted malaria vaccine is currently being implemented in selected regions in Ghana, thanks to better community engagement and trust-building strategies deployed by the vaccine team.

Notwithstanding the vital role clinical trials play in ensuring disease prevention and quality healthcare delivery, lack of trust in the design and implementation of clinical trials could undermine their successful implementation as was the case in the failed EVD vaccine trial in Ghana [5]. Previous studies have reported a number of factors such as apprehensions about drawing and use of blood samples, fear of possible harm and adverse effects of clinical trial medicines negatively affected trust in clinical trials conduct [8,9].

While clinical trials have evolved and improved over time, producing significant advances in diagnosis, treatment and prevention, there are equally key challenges such as issues of transparency and trust [10,11]. The increasing signs of distrust in public institutions and biomedical science clearly suggest a need for examining factors affecting trust in clinical research especially in low-income settings. Therefore, this study explored stakeholders’ views on factors affecting trust in the conduct of clinical trials in Ghana.

## Methods/materials

### Ethical considerations

The study protocol was reviewed and approved by the Ghana Health Service Ethics Review Committee (GHS-ERC007/08/18). Written informed consent was obtained from participants. To ensure confidentiality, codes were assigned to study participants and used on the transcripts.

### Theoretical foundations of the study

Theories help to explain human behaviours and are therefore very important in designing and implementing research that seeks to address public health problem [12]. Thus, Kasperson’s social amplification of risks framework (KSARF) has been adopted and used in the design of this study. Kasperson’s risk framework explains the extent to which a particular risk event merges with psychological, social and cultural procedures to lessen feelings of risk [13]. The framework points out factors that influence risk perception and how risk events are interpreted and communicated by social actors such as institutional stakeholders, traditional and social media in ways that may increase or decrease public reactions to the risk event. According to Kasperson, amplification occurs at two levels: in the transfer of information about the risk, and in the response mechanisms of society. These socially built risk messages are consequently interpreted and acted on by individuals based upon their personal characteristics and attitudes [13]. Social amplification of risk theory is relevant to this work because it provides the framework to analyse individual, community, socio-cultural and geopolitical level actors pertaining to trust in clinical trials and how messages about trial studies are communicated.

### Study design

This was an exploratory study using qualitative research methods of data collection and analysis. In-depth interviews (IDIs) and Key informant interviews (KIIs) were conducted with participants between June and August, 2019. Qualitative research approach helps in capturing feelings, experiences and perceptions of participants on the issue under investigation [14]. Therefore, this approach was considered appropriate in this study because our study aimed at gaining deeper understanding on the role of trust and factors affecting trust in the conduct of clinical trials in Ghana.

### Study site

This was a multi sited-study conducted in the republic of Ghana. Specifically, the interviews were conducted in the Kassena-Nankana East Municipality and Kassena-Nankana West District (Navrongo) of the Upper East Region, Kintampo in the Bono East Region, Hohoe Municipality in the Volta Region and Accra in the Greater Accra region of Ghana.

### Study participants

The study participants included community members (i.e people who have never taken part in clinical trials, those who qualified but have not shown interest to partake in clinical trials, people who were recruited into clinical trials and later dropped out and people who took part in at least 3 clinical trial studies) and community opinion leaders.

Clinical trial scientists who have led clinical trial studies in the three health research centres of the Health Research and Development Division of the Ghana Health Services (i.e the Navrongo Health Research Centre, Kintampo Health Research Centre and Dodowa Health Research Centre), clinical trial monitors and regulatory bodies such as ethics committee members from Navrongo Health Research Centre Institutional Review Board, Ghana Health Service Ethics Review Committee and Ghana Food and Drug Authority members (FDA) were also included in this study. Ghana’s parliament is the legislative arm of government and its mandate include promulgation of laws in addition to other oversight responsibilities. Therefore, the last category of participants in this study comprised members of parliamentary select committee on health.

### Sampling techniques

Purposive sampling method was used to select participants. Purposive sampling is where the researcher selects study participants believed could provide appropriate information to help answer the research questions [15].

### Selection of participants

#### Community and opinion leaders in Navrongo

The Kassena-Nankana East Municipality (KNEM) and Kassena-Nankana West District (KNWD) fall under the research activities of Navrongo Health Research Centre (NHRC). The centre was established in 1989 with the main aim of conducting high quality demographic and health research to inform health policy. NHRC operates the Navrongo Health and Demographic Surveillance System (NHDSS) in the two districts where fieldworkers routinely visit households within the area to collect and update demographic characteristics of the people including their experiences and involvement in clinical trial studies conducted in the area [16]. The list of community members with this experience (i.e those who have participated in at least 3 clinical trials, those who qualified but did not show interest to take part in clinical trials and those who were recruited into clinical trials and later dropped out) was obtained using the NHDSS data base. These individuals were visited by trained data collectors and those who were available and willing to take part in the study were included.

Regarding the selection of opinion leaders, two communities (one in KNEM and the other one in KNWD were selected for the study. Data collectors visited the selected communities and with the support of chiefs and elders, opinion leaders were identified. These opinion leaders were contacted and those who agreed after the purpose of the study was explained to them were interviewed.

### Community members and opinion leaders in Hohoe

First, two communities were selected using purposive sampling method. Data collectors then visited these communities and with the support of sub-chiefs and elders in the two communities, list of some community members and opinion leaders was obtained. These people were contacted and those who agreed were included in the study.

### Clinical trial scientists, monitors, regulators and members of parliament

Various processes were also followed to select these categories of participants into the study. First, official letters were written to the heads of these institutions for their permission to recruit people in their respective institutions into the study. When permission was obtained, the lead author visited these institutions and with the support of the heads, a list of members was obtained. These people were contacted and those who were available and willing to participate in the study were included.

### Training and data collection techniques

Two University graduate research assistants were recruited and trained by the lead author to collect data. In-depth interviews (IDIs) and key informant interviews (KIIs) were the main data collection methods used in this study. The KIIs were conducted with clinical trial scientists, monitors and regulators while the IDIs were conducted with community members, opinion leaders and MPs. Appointments were booked with participants before the interviews were conducted. English language was used to conduct interviews with clinical trial scientists, monitors, regulators and MPs. The interviews with community members and opinion leaders were conducted in the main local languages (i.e Kasem, Nankani and Ewe) spoken in KNWD, KNEM and Hohoe. With consent from participants, all the interviews were audio-recorded using digital voice recorders. A total of 48 interviews (34 IDIs and 14 KIIs) were conducted.

### Data management and analysis

Various steps were followed in the data management and analysis processes. In the first step, recorded interviews were transcribed verbatim after repeatedly listening to them. Two people with previous experience in qualitative research who could understand and speak the three local languages and English were engaged to transcribe the recorded interviews. A codebook containing the main themes and sub-themes was developed by the lead author and reviewed by the second, third and fourth authors. The transcripts were then prepared and imported into QSR Nvivo 12 software to facilitate data coding and analysis. The coding was done by the lead and second authors. The coding process involved a critical review of each transcript and coding of the data into themes. Thematic content analysis was used to analyse the data. The transcripts were prepared and labelled using variables such as age, category of participant and study area. This method did help the study team to compare views on the issues across the different categories during data interpretation. The results were presented as narrative and supported by relevant quotes from the data.

## Results

### Participants’ views on clinical trials conduct

Generally, participants in this study saw the need for the conduct of clinical trials. Clinical trials scientists, monitors and regulators reported that it was important for clinical trials to be conducted to generate new and effective medicines for the management of diseases especially infectious diseases. They added that clinical trials provided useful information on new innovations including vaccines, drugs and medical devices to facilitate the implementation of health policy decisions to improve provision of healthcare services.

> *There is need to conduct clinical trial because we have to provide adequate data on drugs and also to ensure that such drugs are able to treat illnesses they are meant for*. **(KII-clinical trial scientist-07)**
>
> *It is important for clinical trials to be conducted because it is through clinical trials, we are able to improve drug therapies, existing procedures or bring about new innovative procedures to address specific diseases in our country (referring to Ghana). The conduct of clinical trials also helps inform policy decision to improve healthcare management*. **(KII-clinical trial regulator-06)**

some community members and opinion leaders also held that the resistance of microorganisms to some existing medicines (i.e vaccines and drugs) coupled with new emerging diseases, there was the need to conduct clinical trials to find new ways or medicines to address these health problems. Participants added that that since medicines were produced to be used by human beings, it was important for new medicines and other medical devices to be tested on people to help determine the effectiveness of these medicines and devices before they were made available for the general public to use.

> *I think it is good to test them (referring to new medicines) on human beings. If you do not test the medicines, you will not know whether these medicines are good or not*. **(IDI-trial participant-01)**
>
> *Clinical trials are good and I think it is very necessary to conduct clinical trial studies because looking at the situation at hand and you know microorganisms are becoming resistance to a lot of drugs and because there are new emerging diseases, there is need to conduct clinical trials to help address these health problems*. **(IDI-opinion leader-06)**

Some MPs particularly reported that humans were used in other countries to test new medicines and once Ghana belongs to the international community, the country was bound by the principal of reciprocity to also contribute to the development of new medicines and science. As one of them expressed it:

> *We are part of the international community and we benefit from medicines that have been approved through clinical trials conducted elsewhere. Thus, we (referring to people in Ghana) are bound by the principal of reciprocity that if people are used to test such drugs and we are now benefiting from the results of such research work, we also have an obligation to contribute to the development of new drugs in this country*. **(IDI-MP-03)**

Nonetheless, some participants had contrary views about the conduct of clinical trials to test new medicines. These individuals perceived that it was risky for new medicines and other medical devices to be tested on human beings through clinical trial studies. They maintained that an individual could take these trial medicines and develop serious health problems.

> *…Researchers giving me vaccine that is still going through trials, I have a little problem about that; what if after giving me that vaccine and I get serious health problems and die?* **(IDI-community member-03)**
>
> *There are risks in taking trial medicines. Somebody may take these new drugs and die instantly and so there are risks involved*. **(IDI-community member-refused to take part-05)**

Another participant said that taking part in clinical trials, researchers could reveal to people some health conditions, which could affect them psychologically. As one community member put it in the following quote:

> *For me, I do not like clinical trials because they (referring to biomedical researchers) may come and find a particular disease in me, which might rather make me think for the rest of my life*. **(IDI-community member-02)**

### Factors affecting trust in clinical trials conduct

Participants highlighted various factors affecting trust in the conduct of clinical trials. These have been categorized into pre-implementation and implementation factors and discussed below:

### Pre-implementation factors

#### Inadequate community engagement

Participants reported that inadequate community and stakeholder engagement to create awareness on clinical trials affected trust and participation in such studies. Participants believed that where people are not provided with the needed information on rationale of clinical trials through community engagement, it could create doubts and thereby negatively affecting trust in clinical trials conduct. Lack of stakeholders’ engagement and education led to the failed ebola vaccine trial in Ghana according participants.

> *…If you want to conduct clinical trial in Ghana people may want to know about the rationale and benefits of such a trial to them as participants? If such information is not given to people, they may have doubts on what the researchers are going to do and once people are not sure of the need for that trial, they will not want to take part. Lack of information really affected the ebola vaccine trial in Ghana*. **(IDI-MP-02)**
>
> *The researchers did not educate the general public about the malaria vaccine. There was no information such as how the vaccine came about, the side effects and all those things. If you do not have information about these issues, how will you take part?* **(IDI-community member-refused to take part-03)**
>
> Clinical trial monitors and regulators also reported that lack of information could lead to misconceptions about clinical trials conduct such as being used as guinea pigs. According to them, stakeholders mistrust resulting from lack of information contributed to the failed ebola vaccine trials in Ghana.
>
> *…*.*For instance, the Ebola vaccine trial, there was so much noise about it and people who were even educated did not understand what the study was all about. When it happens like that and the information gets out there to the community, everyone will interpret it the way he/she wants and that brings about distrust*. **(KII-clinical trial regulator-03)**
>
> *The problem is that, majority of Ghanaians do not know what clinical trials are all about and that is the underlining fact. People think that biomedical research is all about laboratory work. When people are now invited to participate, then, they say why? You want to use us as guinea pigs?* **(KII-clinical trials monitor-06)**

### Rumours and negative influence

Views expressed by participants in this study suggested that rumours about risks of clinical trials affected trust and participation in such studies. Clinical trial scientists and regulators reported that misinformation given to community members by some individuals or group of people about perceived risks associated with the conduct of clinical trials negatively affected trust in clinical trials conduct. They gave examples of how community members were negatively influenced through audios, videos and WhatsApp messages that were circulated on social media platforms and internet advising people not to take part in the planned ebola vaccine trial and the piloted malarial vaccine in Ghana.

> *Community members were not having problem, but information and feedbacks that were given to them by other people created doubts in their minds about the ebola vaccine trial. For the malaria vaccine, I think everything was ready to go, only for an independent group somewhere I think outside the country, to send WhatsApp messages frightening everybody about the vaccine*. **(KII-clinical trial scientist-03)**
>
> *The current malaria vaccine trial, the problem came from outside the country because some people got videos from a certain lady who circulated them on WhatsApp painting a bad picture about clinical trials. So, it is because of some of these campaigns that is making people to have doubts*. **(KII-clinical trial monitor-05)**

Some community members also recounted instances where negative influence from individuals through audio recordings had influenced their trust and decision to drop out of the piloted malaria vaccine exercise in Ghana. As one of them put it:

> *It was my* spouse *who refused for the child to receive the malaria medicine. My partner actually heard information from audio recording that researchers bring these medicines to test on us and see whether these medicines are good or not. In the audio, it was said that the malaria medicine was tested on human beings and some of them died, so, people should be careful with medicines that researchers bring into the country and ask people to take*. **(IDI-community member-dropped out-04)**

The issue of being used as guinea pigs and side effects were among the propaganda used by family members and friends to influence people negatively about clinical trial studies according to views shared by some opinion leaders.

> *Most at times safety is the main complaint. People who are against clinical trials said it is not safe, it will affect you and that is why researchers will not do it in Europe or America and they want to use our people as guinea pigs*. **(IDI-opinion leader-06)**
>
> *Some educated people are polluting the minds of community members. These educated people will say do not volunteer to take part, don’t you know that there are risks involved? So, whatever the educated person will tell the family members, they will believe it*. **(IDI-opinion leader-02)**

### Implementation factors affecting trust in clinical trials

#### conduct Perceived risks and uncertainties

Some community members held that fear of risks such as side effects, uncertainties about treatment efficacy and in some cases, perceived death that could result from using some clinical trial medicines negatively affected trust and participation in clinical trials. They explained that experience of side effects from routine vaccinations people had received in the past contributed to low trust in clinical trials conduct. They gave examples on how people were scared of the piloted malaria vaccine exercise, which made them refuse their children participation in the exercise.

> *When the malaria vaccine came people were not ready to use it. They said that the researchers are sitting there and collecting vaccines, injecting and killing children*. **(IDI-community member-refused to take part in trial-03)**
>
> *The fear of risks and site effects of trial drugs negatively influences trust of people. If trial participants are not certain of what they are taking, then there is fear and once there is fear, trust is low. If I am not sure of what the trial drug will do to me then, I will not trust and take it*. **(IDI-MP-05)**

It was reported by one opinion leader that people had the perception that using clinical trial drugs could lead to impotency and also shortened an individual life span.

> *The other thing is that some people in our communities have the believe that some of these trial drugs could lead to impotency while others think it could reduce the number of years an individual will live on earth for using trial drugs*. **(IDI-opinion leader-04)**

### Drawing and use of biomedical samples

Drawing of biomedical samples such as blood during clinical trials was reported as another factor affecting trust in clinical trials conduct. According to participants, the inability of clinical trial scientists to explain to community members on reasons for drawing and use of blood samples led to doubts on what exactly these samples were used for. This created avenue for misconceptions by some community members that blood samples were drawn and sold by clinical trial scientist. These misconceptions invariably affected trust and individuals’ interest in clinical trials especially studies that involved drawing of blood samples from participants.

> *Some community members think that researchers take blood samples and sell. There is this woman, we all gave birth at the same period, when the research people came to recruit us into the study, the spouse refused with the reason that researchers will only take blood from study participants and sell for money*. **(IDI-trial participant-05)**
>
> *I have a problem with drawing of blood because you would not know what researchers are going to use the blood for. They (referring to researchers) will just take participants’ blood and they will not tell you what the blood will be used for and you will now be thinking as to what exactly the researchers are going to do with the blood*. **(IDI-community member-dropped out-04)**

The issue of drawing blood samples from clinical trial participants and the perceptions that these samples were being used for spiritual purposes was also reported by some opinion leaders and MPs.

> *Some people normally refuse to give out blood because they belief that the blood is not used only for research purpose*. **(IDI-opinion leader-03)**
>
> *Sometimes, the fear is there and people believe that researchers use these blood samples for juju (referring to spiritual purposes*). *Therefore, if researchers take blood samples from somebody and they do not explain, the person will say oh, no, researchers are taking my blood for spiritual purposes*. **(IDI-MP-04)**

The apprehensions about drawing and use of blood samples and how that affects trust in the conduct of clinical trials was affirmed by clinical trial scientists, monitors and regulators as explained by one of them in the following quote:

> *Drawing of blood samples has been a very big problem and that makes clinical trial very difficult to conduct. The reason is that blood is very significant in our local communities because some people believe that life is in the blood and so, when you take somebody’s blood you have actually come in contact with the person’s spirit. There is also rumour that blood drawn from study participants is sold by researchers*. **(KII-clinical trial scientist-01)**

### Poor informed consent procedures

The views expressed by study participants suggested that poor or inappropriate informed consent administration could affect trust and participation in clinical trial studies. For instance, community members and opinion leaders reported that if clinical trial staff did not properly explain study procedures to potential participants, it could affect trust and their decision to participate in such studies.

> *In an attempt to get many people enrolled into a trial, they (referring to trial team) may not even explain the issues for the people to really understand and actually make an informed decision to be part of the study. Data collectors may give different information to the participants, which may not be in the consent form and that could lead to mistrust*. **(IDI-opinion leader-Hoheo-06)**
>
> *I am wondering whether the fieldworkers do not know the issues or what? Because my spouse asked them questions and they could not answer. They rather asked my spouse to go and see their bosses and that my spouse should just read the consent form and sign it for them*. **(IDI-community member-refused to take part-03)**

Similar sentiments were shared by clinical trial scientists on how poor informed consent administration could significantly affect trust in clinical trials conduct. One of the clinical trial scientists emphatically stated that serious attention was not given to informed consent by clinical trial investigators in terms of how well data collectors were trained to administer informed consent to participants prior to their recruitment into these studies.

> *As for informed consent, it is a huge problem and I do not know whether we should call it informed again. Researchers are supposed to give study information and allow people to ask questions regarding their fears that is not being done. So, informed consent is a big problem affecting trust and I think we (referring to researchers) have not paid much attention to it*. **(KII-clinical trial scientist-02)**

### Perceived no illness

Some community members did not understand why they should use trial medicines although they were not diagnosed to be sick or having a health problem. According to these individuals, it was difficult to trust and take such medicines.

> *I have a problem because I am not sick and yet researchers want to give me medicine! I do not really understand that*. **(IDI-community member-03)**
>
> *I do not like clinical trials because as I am seated, I am very healthy then researchers will come and give me vaccine why?* **(IDI-community member-02)**
>
> *The issue is that I am not sick and yet you (referring to researchers) will bring medicine in the name of helping research, I will not agree*. **(IDI-community member-refused to take part-06)**

## Discussion

This study explored stakeholders’ views on the conduct of clinical trials and factors affecting trust in clinical trials conduct in Ghana. Generally, participants saw the need for clinical trials to be conducted in Ghana. Apart from helping in the advancement of medical science, participants said the conduct of clinical trials enables researchers to come out with new medicines for the management of diseases especially infectious diseases. Participants noted that the resistance of microorganisms to existing medicines and the need to find new medicines to replace old ones necessitates the conduct of clinical trial studies. Medicines are produced to be used by humans and therefore, it is important for these medicines to be tested on human beings to ensure safety and effectiveness of new medicines before they are deployed for public use. The views expressed by participants suggest that conducting clinical trials is necessary to ensure that medicines and other medical devices are safe and would not cause harm when people use them. These findings corroborate previous studies that demonstrated that clinical trials are conducted to determine the efficacy and safety of new medicines and treatments [9,17].

Community perceptions in biomedical research remain critical particularly in low- and middle-income countries [18]. Perception is a constructive process that allows individuals to make inferences about what they see and think about a particular phenomenon, in this case the conduct of clinical trials [19). Across sub-Sahara Africa, evidence suggests that the conduct of clinical trials has largely been perceived to have positive impact on the lives of people especially those who take part in these clinical trials largely because of benefits such as prompt and free medical care [20]. Therefore, the extent to which these clinical trial studies are perceived to be beneficial could be one of the reasons why participants in this study saw the need for the conduct of clinical trials.

Study participants mentioned several pre-implementation and implementation factors affecting trust in the conduct of clinical trials. The findings revealed that inadequate stakeholder engagement to create awareness about biomedical research largely affects trust in their conduct. For instance, where people are not provided with required information on the need for clinical trials to be conducted, they could form their own opinions about such studies. If these perceptions happen to be negative, it could affect their trust in clinical trials conduct. It is not surprising therefore that inadequate stakeholder engagement or education has been reported by participants as one key factor affecting trust in clinical trials conduct. It has been largely reported that lack of knowledge and understanding resulting from poor community engagement negatively affects research activities [21].

To make matters worse, rumours and negative influence from friends and family members about perceived risks associated with clinical trials and the issue of being used as guinea pigs greatly affect trust especially, in low- and meddle-income countries. Misinformation about the rationale for clinical trials conduct could cause fear and doubts in the minds of people leading to their mistrust in biomedical research. As proposed in Kasperson’s social amplification of risk framework, risk events are interpreted and communicated by social actors such as institutional stakeholders as well as social media, and based on the interpretations attached by these communicators, could increase or decrease trust in public health interventions such as clinical trials [13]. These issues were widely reported in this study particularly by clinical trial scientists. These findings also support earlier studies, which demonstrated that rumours about risks in the conduct of clinical trials seriously affected trust and community participation in clinical trial studies [22, 8].

The findings also revealed personal risk and fear as very important factors responsible for low trust in the conduct of clinical trials. This is consistent with previous studies demonstrating that people were not willing to take part in clinical trials because of perceived risks and some cases uncertainty about clinical trial medicines [23]. The current study also found that uncertainty about clinical trial medicines negatively affects trust and decision of people to get involved in such studies. The possibility of new medicines not being effective and the perception that taking medicine that is yet to be proven to be effective was viewed as being experimental in nature and this has been an important factor negatively affecting trust in clinical trials conduct according to views expressed by participants in our study. Other studies both in low and high-income countries have also reported perceived side effects and fear of possible harm as factors affecting trust in clinical trials [24,23]. This means that the issue of fear of risks and its influence on trust in clinical trials conduct is not only a problem in low-and meddle-income countries. Therefore, clearly communicating study procedures, risks and benefits to community members especially potential clinical trial participants and giving them sufficient time to ask questions and have their concerns addressed could improve and maintain trust in clinical trials conduct [25].

Furthermore, the views shared by a good number of participants in this study suggest that people are suspicious that blood samples drawn from clinical trial participants may be sold by clinical trial scientists or being used for other purposes instead of using it for the intended purpose. This generated a mixed sense of anxiety and mistrust in the conduct of clinical trials. The views expressed by both study participants and in literature suggest that the inability of clinical trial scientists to educate community members on reasons for drawing blood samples from participants affects trust leading to doubts on the use of blood samples [26]. Closely related to this is the issue of poor informed consent procedures prior to recruitment of people into clinical trial studies, which has also been described as key factor affecting trust. The inability of data collectors to appropriately administer informed consent to participants during clinical trial studies could largely be as a result of poor or inadequate training. Informed consent administration is viewed as very vital in the conduct of clinical trials and depending on how well clinical trial staff are able to engaged and explain study procedures to potential participants, could positively influence their trust [27]. Clinical trial scientists must ensure that data collectors are adequately trained to build their capacity and knowledge on how to appropriately engage community members and explain clinical trials design and procedures especially potential clinical trial participants during informed consent administration prior to their recruitment into such studies. This is very important not only to help people to take informed decisions, but also improve their trust in clinical trials conduct.

### Study limitations

The major limitation is that since the study used purposive sampling, a non-probability sampling method to select participants to share their opinions on factors affecting trust in clinical trials conduct in Ghana, the views expressed by participants are their personal views and may not necessarily represent views of the larger population.

## Conclusion

The need for biomedical research to be successfully designed and conducted especially in low and middle-income countries where there is high outbreak of infectious diseases is very necessary. However, trust is a fundamental factor affecting a successful conduct of clinical trials as pointed out in our study.

Therefore, there is need for collective efforts by all stakeholders particularly academic institutions, research institutions, Ministry of health, Ghana health services and civil society organizations as well as clinical trial regulatory bodies to take the issue of trust and factors affecting trust in the conduct of clinical trials seriously. In addition, investment in national, regional and community level trust-building activities through appropriate community and stakeholder engagement strategies is highly recommended to address the issue of mistrust resulting from misconceptions people have in clinical trials conduct. Further, improved informed consent procedures through effective training is critical to improving and maintaining trust of community members in clinical trial studies especially in low- and meddle-income settings such as Ghana.

## Data Availability

Portion of the data was used in this study. The data will be made available upon request by the journal

## Acknowledgement

The authors wish to thank all the study participants who shared their views with the study team on the topic. We are also grateful to the research assistants (Mr. Isaac A. Ayaga and Miss. Enyonam Duah) who helped us during data collection.

